# Decline of SARS-CoV-2 specific IgG, IgM and IgA in convalescent COVID-19 patients within 100 days after hospital discharge

**DOI:** 10.1101/2020.08.17.20175950

**Authors:** Huan Ma, Dan Zhao, Weihong Zeng, Yunru Yang, Xiaowen Hu, Peigen Zhou, Jianping Weng, Linzhao Cheng, Xueying Zheng, Tengchuan Jin

## Abstract

Monitoring the levels of SARS-CoV-2 specific antibodies such as IgG, M and A in COVID-19 patient is an alternative method for diagnosing SARS-CoV-2 infection and an simple way to monitor immune responses in convalescent patients and after vaccination. Here, we assessed the levels of SARS-CoV-2 RBD specific antibodies in twenty-seven COVID-19 convalescent patients over 28–99 days after hospital discharge. Almost all patient who had severe or moderate COVID-19 symptoms and a high-level of IgG during the hospitalization showed a significant reduction at revisit. The remaining patients who had a low-level IgG during hospitalization stayed low at revisit. As expected, IgM levels in almost all convalescent patients reduced significantly or stayed low at revisit. The RBD-specific IgA levels were also reduced significantly at revisit. We also attempted to estimate decline rates of virus-specific antibodies using a previously established exponential decay model of antibody kinetics after infection. The predicted days when convalescent patients’ RBD-specific IgG reaches to an undetectable level are approximately 273 days after hospital discharge, while the predicted decay times are 150 days and 108 days for IgM and IgA, respectively. This investigation and report will aid current and future studies to develope SARS-CoV-2 vaccines that are potent and long-lasting.

## Introduction

The 2019 novel coronavirus (later renamed as SARS-CoV-2 in February 2020) infected over 12 million people globally by early July and caused mild to severe COVID-19 patients in millions. Monitoring the levels of antibodies such as immunoglobin (Ig) G, M and A that are specific to SARS-CoV-2 and present in blood provides not only an alternative method for diagnosing SARS-CoV-2 infection (including asymptomatic carriers), but also an simple way to monitor immune responses in convalescent patients or after vaccination. A high and persistent level of SARS-CoV-2 specific antibodies, especially those that can bind to and neutralize the virus, would be a strong indication that an immunized host could resist to SRAS-CoV-2 infection. Currently, there are no effective drugs to specifically prevent or cure SARS-CoV-2 infection; therefore, host immune responses and antibody-based therapeutics will continue playing important roles in combating and later preventing COVID-19.

We previously established a set of diagnostic kits that quantitatively and sensitively measure the levels of serum IgG, IgM and IgA specific to the SARS-CoV-2 spike protein receptor binding domain (RBD), based on a cohort of 87 hospitalized COVID-19 patients and 483 negative controls (Ma et al., 2020). These antibodies specifically bind to the RBD may block its interaction with a cell-surface protein ACE2 that serves as a main viral receptor. Previous studies demonstrated that the serum level of IgG that specifically binds to the RBD highly correlates with that of neutralizing antibody activity in blocking infection of SARS-CoV-2 or a pseudo-virus (Ni et al., 2020; Robbiani et al., 2020; Wu et al., 2020). Our RBD-specific, chemiluminescence-based kits are highly quantitative and sensitive for detecting SARS-CoV-2 elicited IgA, IgG and IgM in blood (Ma et al., 2020). During the optimal detection window of 16-25 days post illness onset, levels of RBD-specific IgA and IgG, but not IgM, were significantly higher in severe and moderate than mild COVID-19 patients (Ma et al., 2020).

## Results and Discussions

To assess the levels of SARS-CoV-2 specific antibodies in COVID-19 convalescent patients over time after hospital discharge, we used the same kits for detecting RBD-specific IgG, IgM and IgA levels in blood of patients in this cohort as we did for them during the hospitalization(Ma et al., 2020). Thirty-three convalescent patients living in Anhui Province of China voluntarily came back to our clinic for revisit, 28–99 days after hospital discharge. Six of them were detected positive for SARS-CoV-2 nucleic acids and excluded in the current serology study. The information of 27 qualified convalescent patients is listed in **Supplemental table 1** in the order of COVID-19 severity during hospitalization. The table includes clinical information, discharge and revisit dates, and interval (28–99 days with a median 91 days) for each patient. The levels of the RBD-specific serum IgG, IgM and IgA (measured as relative light unit or RLU after 40-times dilution) shortly before discharge and at revisit are tabulated at **Supplemental Table 2**. In **Figure 1 (A-C)**, we plotted antibody levels soon before discharge and at revisit as Cutoff Index (COI), which is the ratio of RLU Signal / Cutoff value determined previously for serum IgG, IgM and IgA, respectively(Ma et al., 2020). Among the 27 convalescent patients, all (except #10) who had severe or moderate COVID-19 symptoms and a high-level of IgG during the hospitalization showed a significant reduction at revisit (**A**). The remaining patients who had a low-level IgG during hospitalization stayed low at revisit. As expected, IgM levels in these convalescent patients reduced significantly or stayed low at revisit, except #14 (**B**). The RBD-specific IgA levels were also reduced significantly at revisit (**C**), except patient #10 who also had an increased IgG, but not IgM. Few exceptional cases will need further studies.

**Figure 1.**
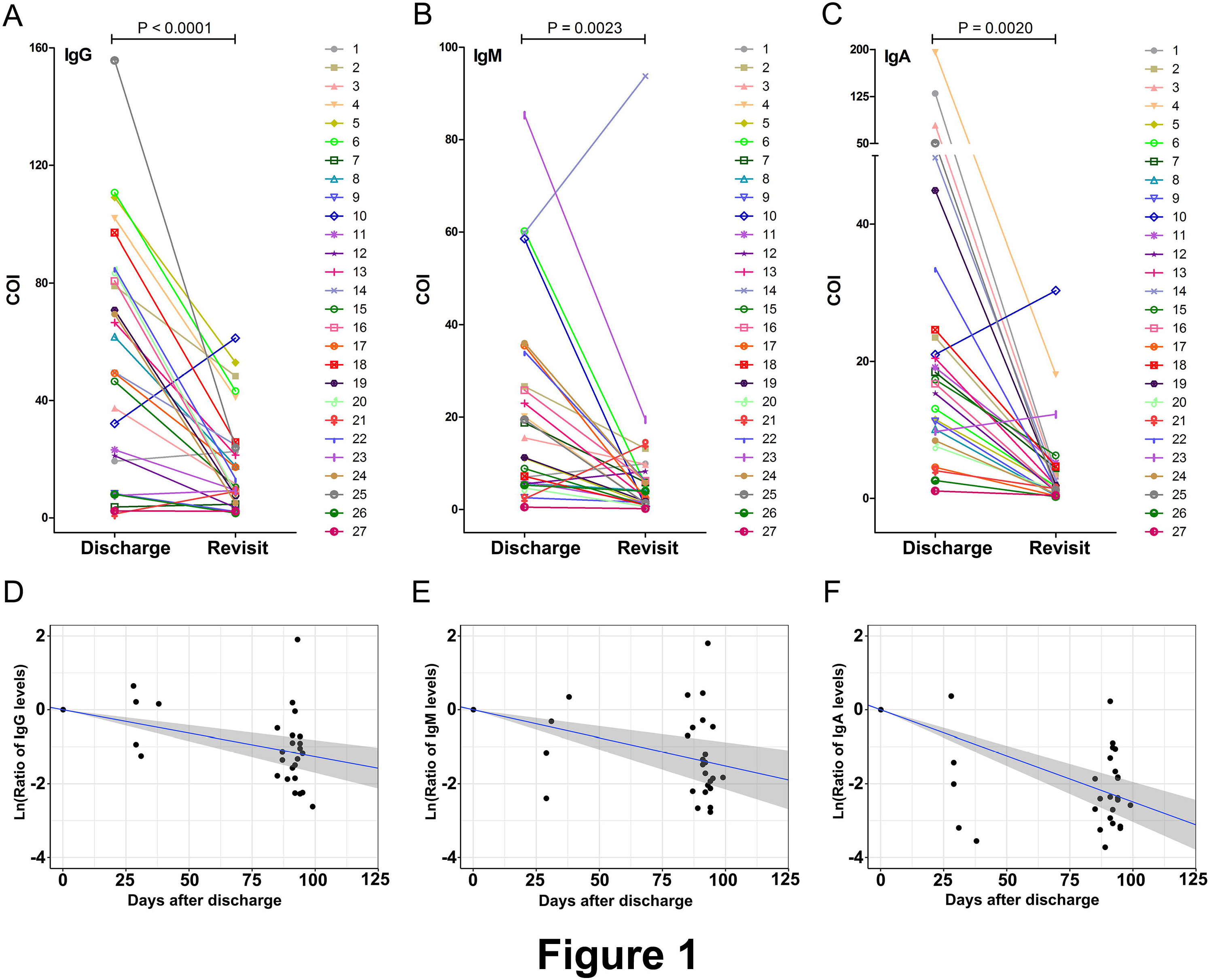
Changes of SARS-CoV-2 RBD-specific serum IgG, IgM and IgA levels in 27 convalescent patients near hospital discharge and at revisit 28–99 days after discharge. (**A-C**) The antibody levels are presented as Cut-Off Index (COI) which is calculated as RLU signal divided by the Cut-Off value previously set for each of IgG, IgM and IgA, respectively. The p values for the difference between discharge and revisit are < 0.0001, 0.0023 and 0.0020 for IgG, IgM and IgA, respectively. (**D-F**) Decline curves for RBD-specific IgG (D), IgM (E) and IgA (F) over time, based on a mathematical model of exponential decay after its peak at recovery (soon before or at discharge). The ratios of COI at revisit versus discharge (day 0) is plotted by log_10_ scale for each patient’s IgG, IgM and IgA separately, as a function of time (days after discharge). See more details in supplemental Methods. The decay curve is marked as a blue line, and 95% confidence interval is marked as a grey zone for each type of SARS-CoV-2 specific antibodies.

We also attempted to estimate decline rates of virus-specific antibodies using a previously established exponential decay model of antibody kinetics after infection(Teunis et al., 2016). Based on the combined data of COI ratios before and after discharge for each of the 27 patients, we plotted decay curves for RBD-specific IgG, IgM and IgA over time (**Figure 1D–F**). The predicted days when convalescent patients’ RBD-specific IgG reaches to an undetectable level are approximately 273 days (ranging from 134–304 days or 4.5–10 months) after hospital discharge (D), while the predicted decay times are 150 days and 108 days for IgM and IgA, respectively.

In summary, the initial data of this longitudinal study showed that the levels of SARS-CoV-2 RBD-specific antibodies in most COVID-19 convalescent patients reduced significantly or remained low within the first 100 days after discharge. A mathematical modeling and extrapolation predicts that the virus-specific IgG in this group of convalescent patients will disappear in 273 days (∼ 9 months). Our data and analyses provide timely and critical information on how long acquired humoral immune responses to this new human coronavirus could persist. In literature, there are so far few papers on the persistence of the SARS-CoV-2 elicited antibodies after recovery beyond two weeks(Long et al., 2020; Ni et al., 2020; Robbiani et al., 2020; Wu et al., 2020). In one study, blood samples (both cells and plasma) of six convalescent patients were collected two weeks after discharge and used to examine humoral and cellular immune responses(Ni et al., 2020). In another study (Wu et al., 2020), the serum IgG specific to the SARS-CoV-2 RBD and virus-neutralizing antibodies remained similarly low in 47 recovered patients two weeks after discharge. However, a recent study reported drastic declines of RBD-specific IgG and virus-neutralizing activities in 148 convalescent patients after an average of 39 days (Robbiani et al, 2020). The most recent study reported that 12.9% of the symptomatic group and 40% of the asymptomatic group became negative for IgG after 8 weeks, consistent with our findings of up to 99 days or 14 weeks.

Our current and other related studies point out a conclusion that SARS-CoV-2 infection did not elicit a long-last humoral immune memory. Similar to what reported with the SARS-CoV-1 infection (Cao et al., 2007). Our observation and decline kinetics modeling provide a guideline for SARS-CoV-2 vaccine designing as how to achieve long-last humoral immune response and memory. One way is to seek immunogens and adjuvants that show very strong immune responses such as virus-specific IgG induction that can be easily monitored. For example, a recent clinical trial showed that an experimental vaccine using inactivated SARS-CoV-2 viruses with alum as the adjuvant only elicited comparable to, but not much higher virus-specific IgG production than what we and others observed in hospitalized COVID-19 patients (Xia et al., 2020). Using more potent immunogens and adjuvants to enhance immune responses for stronger SARS-CoV-2 IgG production will be an important early indication for effective development of SARS-CoV-2 vaccines that are highly potent and long-lasting. Although long-term data beyond 99 days after discharge are still in progress and needed to confirm our modeling, our current report provides timely information and fills the gap of knowledge to assess the persistence of antibody levels in response to this novel human coronavirus. A rapid reduction of antibodies (IgG, IgM and IgA) specific to SARS-CoV-2 we observed in convalescent patients examined 4–14 weeks after discharge warrants timely and close attention; however, one shall interpret our current data cautiously. First, we had data so far from a relatively small group of COVID-19 convalescent patients, who were first chosen because we can compare changes of the virus-specific antibodies after discharge. Second, we measured only the antibodies specific to SARS-CoV-2 RBD in the study subjects. Third, we have not examined cellular immune responses in this cohort of patients as did by others (Grifoni et al., 2020; Ni et al., 2020). Overall, our data are similar to what reported with SARS-CoV-1 infection: patients recovered from SARS had a rapid IgG decline and became undetectable after 3 years (Cao et al., 2007). However, a study reported the presence of long-lasting memory T cells reactive to the SARS-CoV-1 N protein in SARS patients recovered from 17 years ago (Le Bert et al., 2020). Nonetheless, our observational and longitudinal serology study provides timely and valuable information to aid current and future studies, to address important issues such as how to use convalescent plasma or hyper-immunoglobins to treat COVID-19 patients and how to develop SARS-CoV-2 vaccines that are highly potent and long-lasting.

## Data Availability

I promise that all data in this research are true and reliable

## Funding

T.J. is supported by the Strategic Priority Research Program of the Chinese Academy of Sciences (XDB29030104), National Natural Science Fund (Grant No.: 31870731 and U1732109), the Fundamental Research Funds for the Central Universities (WK2070000108). TJ is supported by a COVID-19 special task grant supported by Chinese Academy of Science Clinical Research Hospital (Hefei) with Grant No. YD2070002017. M.H. is supported by the new medical science fund of USTC (WK2070000130). JW and XY are supported by the Fundamental Research Funds for the Central Universities with Grant No. YD9110004001 and YD9110002002, respectively.

## Acknowledgements

We thank the staff and patients in The First Affiliated Hospital of USTC for their support in providing samples and clinical data collection. We would also like to thank Profs. Tian Xue and other colleagues in Division of Life Sciences and Medicine for their generous and professional support.

## Conflict of Interest Disclosures

Authors declare that they have no conflicts of interest.

## Notes

### Competing Interest Statement

The authors have declared no competing interest.

### Author Declarations

This study was reviewed and approved by the Medical Ethical Committee of First Affiliated Hospital of USTC (approval number 2020-XG(H)-014 and 2020-XG(H)-009).

